# Prognostic value of serum MMP-9 level in acute ischemic stroke patient

**DOI:** 10.1101/2022.06.01.22275560

**Authors:** Amit R. Nayak, Swapnil S. Muley, Rajpal S. Kashyap, Neha H. Lande, Dinesh P. Kabra, Lokendra R. Singh, Hatim F. Daginawala

## Abstract

**Purpose:** MMP-9 is reported to be a marker of severity and worse outcome after stroke. Despite the above information, there are very few studies based on the evaluation of role of follow-up estimation of serum MMP-9 in acute ischemic stroke (AIS) patients. The purpose of this study was to evaluate the role of follow-up estimation of serum MMP-9 in the prognosis of AIS patients. MMP-9 levels were also compared with reported biomarkers viz. Neuron-Specific Enolase (NSE), Glial specific protein (S100ββ) and Inter-α-trypsin inhibitor heavy chain 4 (ITIH4).

**Materials/methods:** MMP-9, NSE, and S-100ββ were measured in serum samples collected at admission and expired/discharge time of AIS patients (n=48) using commercially available ELISA kits. ITIH4 was measured using Indirect ELISA protocol.

**Results:** MMP-9 at admission (3639 ±1471) and discharged/expired (3707 ±1268) was significantly higher (p<0.05) in AIS patients as compared to healthy control subjects (749 ±302). Expired patients showed significantly (p<0.05) higher MMP-9 as compared to survivors. Similarly, follow-up MMP-9 at discharge was higher in AIS patients who expired within one year (75%; 3/4). The observed values of MMP-9 significantly correlates (p<0.05) with NSE (r=0.452), S-100ββ (r=0.452) and ITIH4 (r=-0.2027).

**Conclusions:** Follow-up measurement of serum MMP-9 may help to predict both short term and long term mortality in AIS patients.

## Introduction

Brain Stroke is continued to be a measure health concern worldwide [1]. Acute ischemic stroke (AIS) is most common form of cerebrovascular disease and account for approximately 85% of total incidences [2]. The alarming fact about AIS is that currently, no specific neuroprotective treatment is available for AIS. Till today intravenous tissue plasminogen activator (IV-tPA) is the only available, FDA approved treatment for AIS. However, use of IV-tPA for the treatment is very limited, because as per the guideline tPA treatment can be given to stroke patient if the patient reached hospital within the window period maximum 4.5 hour [3,4]. The complications do not end here, as it is accompanied by a more lethal phenomenon known as cerebral edema in few cases [5]. Therefore, there is need for a biomarker, which will provide satisfactory information on injury mechanism, helps to monitor progress of the disease and response to the treatment.

Matrix metalloproteinases are zinc and calcium-dependent endopeptidases that are capable of degrading all components of the ECM [6]. Twenty three MMPs have been identified till date, of which MMP-9 is most widely studied in acute ischemic stroke (AIS) [7-9]. MMP-9 expression was reported to increases in the rat’s brain after 3 of transient ischemia **and reperfusion [10]**. Experimental studies have shown that MMP-9 knock-out mice were protected against ischemic and traumatic brain injury by reducing proteolytic degradation of critical blood-brain barrier and white matter components [11,12]. Similarly, increase MMP-9 levels have also been reported to be associated with cerebral edema development following experimental acute cerebral infarction in rats [13].

It is also been reported as an adjunct marker to CT scanning for diagnosis of stroke patients in the emergency setting [14]. Circulating MMP-9 level is reported to correlates with the National Institute of Health Stroke Scale (NIHSS) used for stroke assessment of severity in stroke patient [15]. Serum MMP-9 Level also predicts hemorrhagic transformation in stroke patient following thrombolytic treatment [16].

In the present study we measured MMP-9 level in the paired blood samples collected at admission & Discharge/Expired time to investigate its prognosis significance. In addition, MMP-9 was also compared with known marker Neuron specific enolase (NSE), S-100ββ and Inter alpha trypsin inhibitor heavy chain 4 (ITIH4).

## 2. Materials and Methods

### 2.1. Study subjects and clinical diagnosis

Fifty five consecutive patients admitted to Central India Institute of Medical Sciences (CIIMS), Nagpur, India within 24h of onset of AIS symptoms were included in the present study. Patient were diagnosis was based on WHO definition of stroke, “rapidly developing signs of focal (or global) disturbance of cerebral function lasting more than 24 hrs (unless interrupted by surgery or death), with no apparent non vascular cause, history, neurological examination and computerized tomography (CT) or MRI”.

Patients with transient ischemic attack, hemorrhage, malignancies were excluded from the study. Other exclusion criteria included post-seizure neurological deficit, brain abscess, tumour, laboratory findings of CNS and other infections (pneumonia, urinary tract, other site), myocardial infarction, surgery, trauma in previous 30 days and specified co-morbidities (end-stage renal/hepatic disease and metastatic malignancy).

Nine age and sex matched individuals without any history of stroke, and carotid stenosis were selected as control.

Neurological deficit and outcome in were assessed using National Institute of Health Stroke Scale (NIHSS) score. All patients were admitted to Intensive Care Unit (ICU), where ambient temperature was between 20 and 25°C. All patients were given standard treatment as per signs symptoms and history. Five patients admitted within window period were thrombolyzed using IV-tPA. Similarly, decompressive hemicraniectomy and duroplasty was done in four patients for malignant middle cerebral artery syndrome. Other treatment regime included antiplatelet agents (aspirin 150 mg and clopidrigel 75 mg once a day), antiedema measures, such as mannitol 20%, 0.25–0.5 g/kg over 20 min (not exceeding a total of 2 g/1 kg of body weight in 24 h), with symptoms of raised intracranial pressure, oral glycerol in addition to mannitol, IV fluids, and other supportive measures, including treatment of concurrent illnesses like hypertension and diabetes mellitus. The protocol for this study was reviewed and approved by Institutional Ethics Committee of Central India Institute of Medical Sciences.

### 2.2. Sample collection

Serum samples from AIS patients were collected immediately at the time of admission and at discharge / expired time. Serum samples from age and sex matched healthy individuals were also taken as control group. All the samples for analysis were stored at -80°C until use.

### 2.3. Estimation of serum MMP-9, NSE and S-100ββ **in AIS patients**

#### 2.3.1. MMP-9 Estimation

The concentration of MMP-9 was measured in serum samples collected at the time of admission and expired/discharge of AIS patients using commercially available ELISA kit (Ray Biotech Human MMP-9 kit). In brief, all reagents and samples were brought to room temperature (18 - 25°C) before use. 100 µl of each standard and sample were added into appropriate wells. Wells were covered with lid and incubated for 2.5 hours at room temperature with gentle shaking. Remaining solution was aspirated and wells were washed 4 times with 1x wash solution. After last wash, any remaining wash buffer was decanted by inverting and blotting the plate against clean paper towels. 100 µl of 1x prepared biotinylated antibody was added to each well and the plate was incubated for 1 hour at room temperature with gentle shaking. Remaining solution was discarded and the plate was again washed 4 times with 1x wash solution. 100 µl of prepared Streptavidin solution was added to each well and the plate was incubated for 45 minutes at room temperature with gentle shaking. Remaining solution was discarded and the plate was washed 4 times with 1x wash solution. 100 µl of TMB One-Step substrate reagent was added to each well and incubated for 30 minutes at room temperature in the dark with gentle shaking. Finally 50 µl of stop solution was added to each well and absorbance was read at 450nm immediately. The minimum detectable concentration using this kit was 10pg/ml.

#### 2.3.2. NSE Estimation

Serum levels were estimated with Can Ag NSE EIA (Sweden) as per the instructions of kit manufacturer. The assay is based on solid phase, non-competitive immunoassay based on two monoclonal antibodies (derived from mice) directed against two separate antigenic determinants of NSE molecule. The Monoclonal Antibodies (MAb) used bind to γ subunit of enzyme and thereby detect both γγ and αγ isoenzymes of NSE. In brief, the method is to transfer the required number of microplate strips to a strip frame. Each strip was washed once with wash solution. 25 µL of the NSE Calibrators (CAL A, B, C, D, and E) along with patient’s specimens (unknowns-Uk) was pipetted into the strip wells. The plate was incubated for 1 hour (±10 min) at room temperature (20-25°C) with constant shaking of the plate using a microplate shaker. After incubation each strip was aspirated and washed 6 times.100 µL of TMB HRP-substrate was added to each well and incubated for 30 mins (± 5 min) at room temperature with constant shaking. After incubation, 100 µL of stop solution was added, mixed and the absorbance was read at 405 nm in a microplate spectrophotometer within 15 min after addition of stop solution.

#### 2.3.3. S-100ββ Estimation

In-vitro assay for the quantitative determination of S-100ββ in human serum was performed as per the instructions indicated in the manual (Can Ag S-100 EIA Sweden). This is a two-step enzyme immunometric assay based on two monoclonal antibodies derived from mouse specific for two different epitopes of S-100ββ. Thus, the assay determines both S-100αβ and S-100ββ without cross-reactivity with other forms of S-100. In brief, ELISA wells were washed once with wash solution. 50 µL of S100ββ calibrators and samples were pipetted into the strip wells. 100 µL Biotin Anti-S100ββ was added to each well and the frame was incubated containing the strips for 2 hours (± 10 min) at room temperature (20–25°C) with constant shaking of the plate using a microplate shaker. After incubation, each strip was aspirated and washed 3 times using the wash buffer. 100µL of Tracer working solution was added to each well. The frame was incubated for 1 hour (± 5 min) at room temperature with constant shaking. After incubation, each strip was aspirated and washed 6 times. 100µL of TMB HRP-Substrate was added to each well and incubated for 30 min (± 5 min) at room temperature with constant shaking. 100µL of stop solution was added and mixed. Absorbance was recorded at 405nm in a microplate spectrophotometer within 15 min after addition of stop solution.

#### 2.3.4. ITIH4 Estimation

ITIH4 was estimated using indirect enzyme-linked immunosorbant assays (ELISA). In brief, 100 μl (1:400) serum samples taken from stroke patients were added to individual micro titer wells and blocked with 200 μl of 2.5% BSA in phosphate buffer saline (PBS) for 2hrs. After washing with PBS, the polyclonal antibody against ITIH4 protein was added, and plates were incubated at 37 °C for 45 min. The wells were washed, followed by addition of the secondary antibody (goat anti rabbit immunoglobulin horseradish peroxidase; IgG-HRP) and incubation for 45 min at 37 °C. After another wash, antibody reactivity was detected via the addition of 100 μl Tetramethylbenzidine hydrogen peroxide (TMB/H2O2) substrate solution to the wells, which were then incubated at room temperature for about 5 min. The reaction was stopped with 100 μl of 2.5 N H2SO4, and the absorbance of each well was read at 450 nm.

### 2.4. Statistical analysis

All the statistical analysis was performed using MedCalc (version10). Association of various risk factors was evaluated using chi square test on proportion of patients who belonged to any of the three MMP-9 categories. Similarly, mean of MMP-9 at admission and discharge were compared at risk factor level using t-test. Correlation of MMP-9 with severity and outcome score (i.e. NIHSS), age, NSE and S-100 were calculated using Pearson correlation coefficient. P-value <0.05 were considered as statistically significant.

## 3. Results

Out of 55 AIS patients, 7 were excluded i.e. 5 due to discharge against medical advice while 2 due less sample volume available for test. Remaining 48 AIS patients were included in the study. Three AIS patients expired within 72h of hospitalization were included in “non-survivor” AIS patients group. Forty-five patients who partially improved were discharged from the hospital. Twenty-six discharged patients for whom contact numbers were available were then followed telephonically after one year. Out of 26 cases, four had expired within one year. These patients were included in “Follow-up Expired” group. Twenty-two patients who remained alive till our follow-up period were included in “Follow-up survivor” group. Nineteen discharged AIS cases, for which follow-up status is not available, were included in “loss of follow-up group (Figure 1).

**Figure 1:**
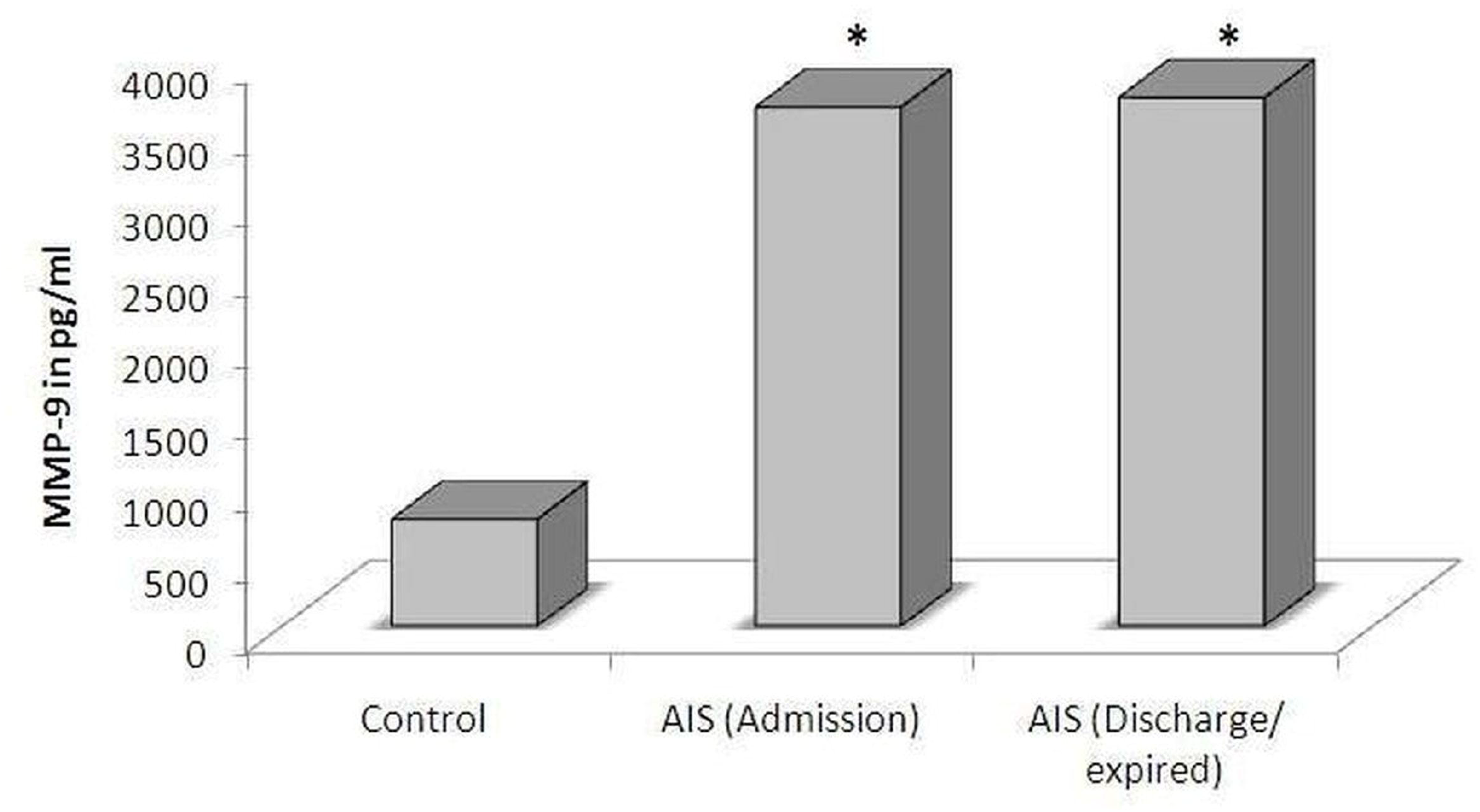
Flow Chart of the study.

MMP-9 levels were estimated in all 45 AIS patients serum samples collected at admission and at discharged/expired time respectively. We found that mean MMP-9 at admission (3639±1471) and at discharge/expired time (3707±1268) was significantly higher (p<0.05) in AIS patients as compared to healthy individual controls (749±302) as shown in Figure 2.

**Figure 2:**
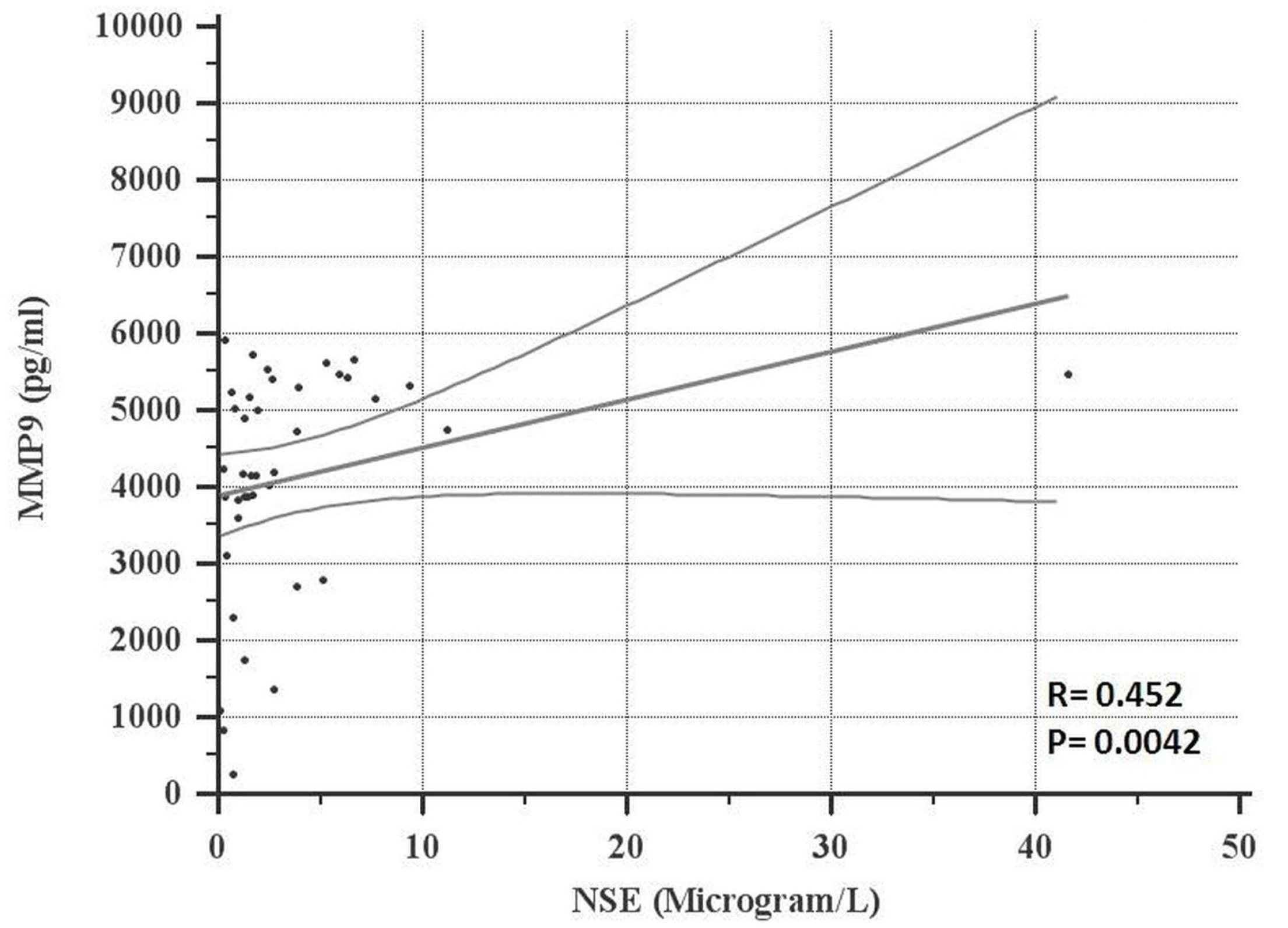
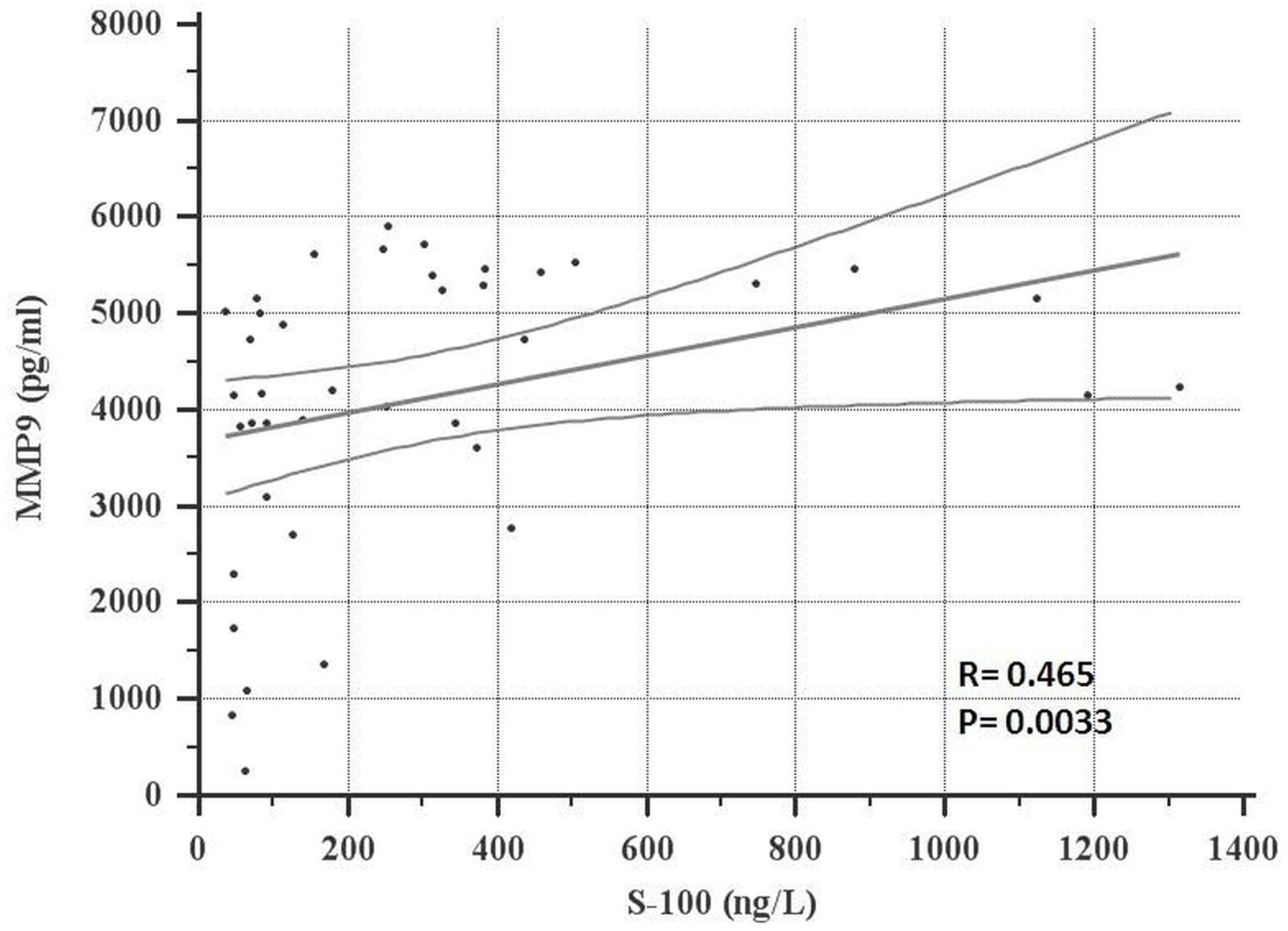
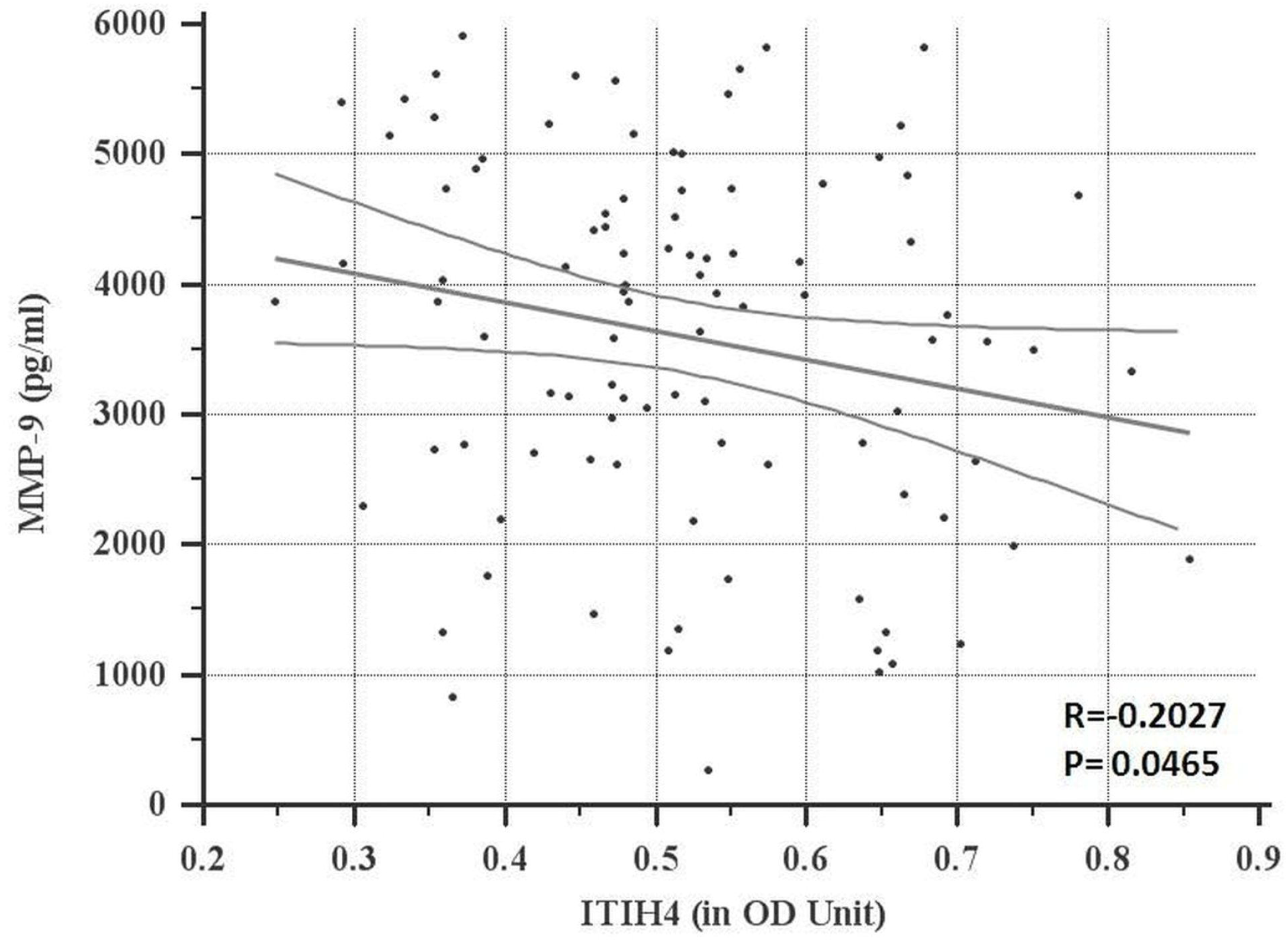
MMP-9 level in AIS patients (n=48) and healthy control subjects (n=9).

MMP-9 levels at admission and discharge/expired time in samples of AIS patients at characteristics level are given in Table 1. We found that AIS patients with smoking habit (vs. nonsmoker), alcoholic habit (non-alcoholic), past history of stroke (vs. fresh cases), AIS patients with Rt. MCA infarct and those who expired (p<0.05 vs. survivor), showed high MMP-9 levels both at admission and at discharge/expire time. Similarly, in follow-up increase in MMP-9 level at discharge time was observed in AIS patients with higher NIHSS score (≥10), multiple ischemic infarcts, and those underwent decompressive surgery and who expired within a year (i.e. Follow-up Expired AIS patients).

**Table 1:** Shows MMP-9 level in admission and discharge/Expired sample of AIS patient (n=48) at various characteristic levels.

| Characteristics |  | n (%) | Admission | Discharge |
| --- | --- | --- | --- | --- |
| Age | $\leq 50$ | 19 (40) | 3714±1630 | 4027±1063 |
| | $\geq 50$ | 29 (60) | 3517±1351 | 3436±1332 |
| Sex | Male | 33 (69) | 3620±1493 | 3612±1200 |
| | Female | 15 (31) | 3539±1414 | 3797±1403 |
| Hospitalization period | $\leq 10$ days | 41 (85) | 3675±1419 | 3614±1212 |
| | $\geq 10$ days | 7 (15) | 3703±1682 | 3995±1548 |
| Smoking Habit | NO | 44(92) | 3492±1460 | 3561±1239 |
| | Yes | 4(8) | 4727±837 | 4863±710 |
| Alcoholic | NO | 41(85) | 3507±1457 | 3576±1290 |
| | Yes | 7(15) | 4113±1431 | 4219±911 |
| NIHSS Score | $\leq 10$ | 21 (44) | 35702±1536 | 3275±1234 |
| | $\geq 10$ | 17 (35) | 3703±1719 | 4087±1428 |
| | ND | 10 (21) | 3466±710 | 3791±714 |
| Thrombolysis | NO | 43 (90) | 3583±1349 | 3577±1246 |
| | Yes | 5 (10) | 3702±2392 | 4469±1148 |
| Decompressive surgery | NO | 44 (92) | 3567±1393 | 3570±1255 |
| | Yes | 4 (8) | 5048±902 | 4111±1560 |
| Stroke and seizure | NO | 45 (94) | 3557±1451 | 3641±1288 |
| | Yes | 3 (6) | 4176±1689 | 4099±481 |
| History of heart disease | NO | 45 | 3651±14565 | 3650±1273 |
| | Yes | 3 | 2765±1405 | 3981±1111 |
| Hypertension | NO | 18 (37) | 3787±1804 | 3869±1253 |
| | Yes | 30 (63) | 3480±1217 | 3550±1262 |
| DM | NO | 36 (75) | 3776±1445 | 3761±1318 |
| | Yes | 12 (25) | 3053±1402 | 3395±1045 |
| Infarct type | Lt. MCA | 14 (29) | 3925±1371 | 3905±1100 |
| | Rt. MCA Infarct | 10 (21) | 4005±1794 | 4242±1023 |
| | Multi-infarct patient | 8 (17) | 3082±759 | 4112±1401<sup>@</sup> |
| | Pontline stroke | 5 (10) | 2170±988 | 2281±1212 |
| | Other | 11 (23) | 3828±1483 | 3161±1058 |
| Outcome <sup>#</sup> | Non survivor (expired within 72hrs.) | 3 (6) | 4617±1606 | 5259±358* |
| | Follow-up Expired (Long term expired) | 4 (8) | 2979±1651 | 3945±909 |
| | Follow-up survivor | 22 (46) | 3339±1640 | 3495±1259 |

Note: <sup>#</sup> indicates Data of 29 patient after excluding loss of follow-up (n=19); \* P- value <0.05 vs Survivor; <sup>@</sup> P- value <0.05 vs Admission concentration

Based on the value of serum MMP-9 in paired samples (i.e. at admission and discharge/expired samples) AIS patients were further grouped into two categories; category 1 (Admission MMP-9 ≥ Discharge MMP-9), category 2 (Admission MMP-9 < Discharge MMP-9). **Table 2** depicts the association of MMP-9 categories with baseline and clinical characteristics of AIS patients. We found AIS patients with smoking habits (P<0.05) and alcoholic habits (P<0.05) show stable or decrease in MMP-9 value at discharge time (Category1). Similarly, diabetes, infarct type and clinical outcomes in AIS patients were found to be significantly (p<0.05) associated with the increase in MMP-9 level at discharge time (Category 2). Among the infarct type 75% (6/8) of multiple infarct and 60% of pontine infarct patients show increase in MMP-9 level at discharge. Likewise 75% (3/4) long-term follow-up expired AIS patients also show increase in MMP-9 level at discharge.

**Table 2:**
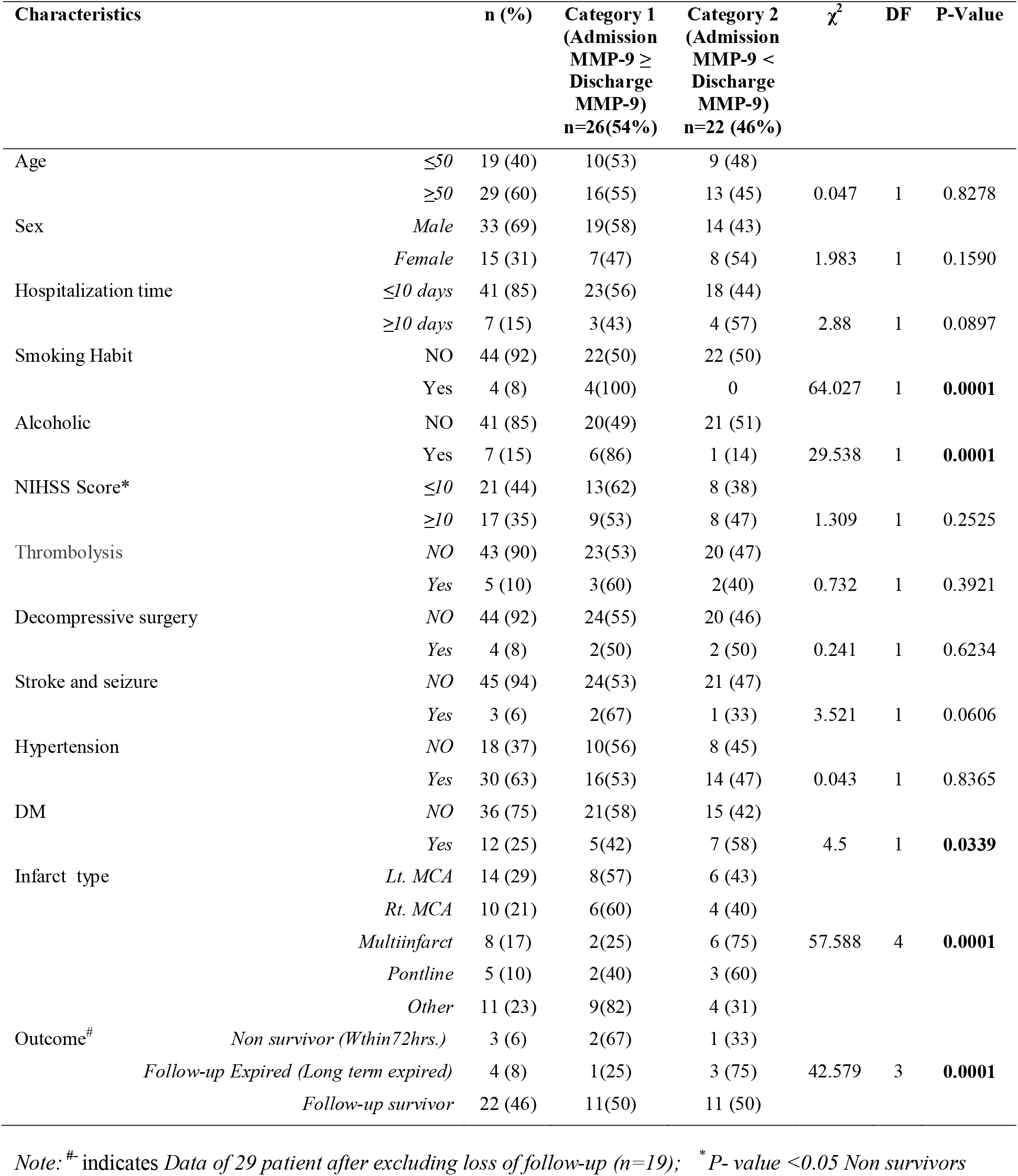
Association of characteristic of AIS patient with different MMP-9 categories

We also correlated MMP-9 level with other established biomarkers (i.e. NSE; S-100ββ and ITIH4). We found that the MMP-9 level positively and significantly correlates with NSE (r=0.452; Figure 3a) and S-100ββ (r=0.452; Figure 3b), while it shows a negative correlation with ITIH4 (r= -0.2027; Figure 3c)

**Figure 3:**
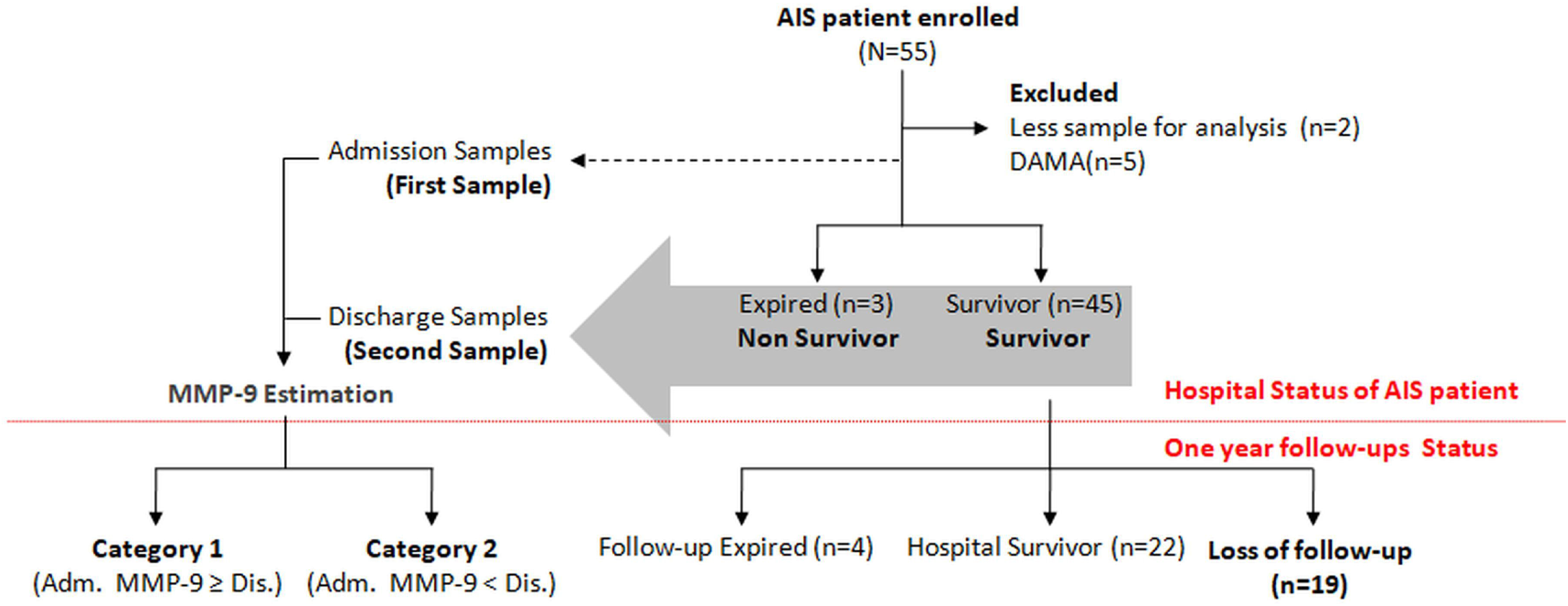
Correlation of MMP-9 (pg/ml) with reported biomarker of AIS, (a) Neuron Specific Enolase (NSE), (b) S-100ββ and (c) ITIH4 in AIS patients.

## 4. Discussion

Early diagnosis and prognosis of AIS is critically important for identifying the risk and planning of better care & treatment of stroke patient towards the better outcome. In the present study investigated the association between serum MMP-9 levels and short term and long term mortality in following ischemic stroke. We found that MMP-9 level was high in AIS patients as compared to healthy subjects. Among the AIS patients, expired patient had a higher MMP-9 level as compared to those who survived. Interestingly, 75% of AIS patients who expired within a year also showed a marked increase in MMP-9 at discharge as compared to its admission value.

MMP-9 is one of the most widely studied inflammatory molecules in AIS patients. MMP-9 activity has been reported to increase in brain tissue and serum after AIS [8,10]. Recently it was reported that MMP-9 mRNA concentration in expired AIS patients was three-fold higher as compared to those who survive, suggesting that estimation of MMP-9 mRNA concentration could predict the outcome in stroke [11].

Earlier females have been reported to be independently associated with a higher risk of poor outcome after stroke [12]. Hoda et al. had reported that MMP-9 expression upregulates in both males and females after stroke and minocycline treatment reduces MMP-9 expression in both genders [17]. Corresponding to the earlier report by Hoda MN, we also noted that MMP-9 expression did not differ in males and females. Similarly, we did not find any significant association of MMP-9 with age and hospital stay of AIS patients.

Lipid-soluble cigarette smoking particles are reported to induce the expression of inflammatory genes in the animal model [14]. Likewise, a high MMP-9 concentration was also reported in sera of alcoholic subjects [15]. We also found high MMP-9 in AIS patients with smoking and alcoholic habit. However, we did not find any follow-up increase in MMP-9 in smokers (category1) while only 14% of alcoholic AIS patients showed follow-up increase in MMP-9. This indicates that although smoking and alcoholic habits might be important risk factors adding to the severity of AIS, they are however less important for the mortality of these patients.

NIHS score which is calculated based on the physical response of AIS patient was used for the assessment of the neurological deficit and outcome in AIS patients [16]. We did not find any major difference in MMP-9 level among AIS patients with <10 or >10 NIHS score at the time of admission, while follow-up increase in MMP-9 at discharge samples was observed in AIS patients who had higher NIHS score (≥10).This further confirms the prognostic importance of evaluation of follow-up MMP-9 estimation.

MMP-9 has also been reported to be a predictor of failure in recanalisation of the artery after thrombolytic or hemorrhagic transformation in AIS patients treated with t-PA [18, 19]. It was also reported that tPA upregulates the expression of MMP-9 in the brain after stroke [20]. Our observation also indicated raised MMP-9 in 40% AIS patients who underwent thrombolysis. A similar observation was also noted in non-thrombolyzed patients (i.e. in 47%). Interestingly, it is noted that two (out of five) thrombolyzed patients with very high MMP-9 at admission expired within 72hrs while two patients who expired after discharge, were associated with secondary complications (patient 1: Basilary artery stenosis and patient 2: diabetics, LV dysfunction with arterial fibrillation, hypothyroidism, and asthma). One patient who did not have high MMP-9 level and any secondary complications have survived during our follow-up after discharge. This suggests that it is very important to check MMP-9 level before thrombolysis. It also suggests that MMPs inhibitor along with tPA may be used in combination for the better thrombolytic response.

Experimental studies using mice model show that MMP-9 is also associated with compromised recovery in diabetic db/db mouse following a stroke [21,22]. Similarly Planas et al. reported high MMP-9 expression in hypertensive patients irrespective of stroke subtype [23]. We also observed a significant association of MMP-9 with diabetic stroke patients while we did not find any association of MMP-9 with hypertension. We also studied MMP-9 level in various infarct conditions. We found higher MMP-9 level among the AIS patients with Rt. MCA infarct. A follow-up increase in MMP-9 was also observed only in AIS patients with multiple infarcts. Earlier Montaner et al. studied the MMP-9 level in AIS patients and reported that MMP-9 levels are associated with neurological deficit, middle cerebral artery occlusion, and infarct volume [24]. Similarly Kaste et al. reported that left MCA occlusion is more fetal than the right MCA [25]. We also observed higher NIHSS score of 14±10 among the AIS patients with left MCA infarct as compared to NIHSS score of 10±10 in AIS patients with right MCA infarct (Data not shown). But in contrast to higher NIHSS score among AIS patients with left MCA infarct, we got higher MMP-9 levels in AIS patients with right MCA infarct. However, this observation corresponds with the report by Finks et al. which suggests that AIS patients with right MCA infarct may have low NIHSS score despite substantial lesion as detected by him in MRI study of AIS patients [26]. This suggests that estimation of circulating MMP-9 level could be a very useful biomarker for accurate diagnosis and prognosis of AIS patients.

We also estimated NSE and S-100ββ and ITIH4 in the same sets of AIS samples used for MMP-9 estimation. Both NSE and S100ββ have been reported to be important markers for predicting short-term functional outcome in AIS patients [27, 28]. Similarly, in our earlier studies we had reported serum ITIH4 as a novel biomarker for prognosis of AIS [29, 30]. We reported that low serum ITIH4 level could be a marker of AIS and its reappearance could be correlated with the improvement in AIS patient [30]. In the present study, we found that the expression of MMP-9 positively correlates with NSE and S-100ββ. Similarly, it also showed a negative correlation with ITIH4. This further confirms that MMP-9 could be a potential candidate marker for prediction of outcome in AIS patients.

Although, the current study has a limitation of having a small sample size, the results obtained are indeed quite interesting. Therefore, further studies are needed to validate the findings of this preliminary study.

In conclusion, the current study suggests that follow-up measurement of MMP-9 could be very useful in the prediction of severity as well as short term and long term mortality in AIS patients.

## Data Availability

All data produced in the present work are contained in the manuscript

## Acknowledgement

All authors would like to acknowledge Dr. Shweta R. Badar & Ms. Anuja P. Kawle for her assistance in statistically analyzing the data & Language editing respectively.

## References

1. GBD 2019 Stroke Collaborators.Global, regional, and national burden of stroke and its risk factors, 1990-2019: a systematic analysis for the Global Burden of Disease Study 2019. Lancet Neurol. 2021 Oct;20(10):795–820.

2. Musuka TD, Wilton SB, Traboulsi M, Hill MD. Diagnosis and management of acute ischemic stroke: speed is critical. CMAJ. 2015;187(12):887–893.

3. Emberson J, Lees KR, Lyden P, et al. Effect of treatment delay, age, and stroke severity on the effects of intravenous thrombolysis with alteplase for acute ischaemic stroke: a meta-analysis of individual patient data from randomised trials. Lancet 2014;384:1929– 35.

4. Kleindorfer D, Kissela B, Schneider A, et al. Neuroscience Institute. Eligibility for recombinant tissue plasminogen activator in acute ischemic stroke: a population-based study. Stroke 2004;35:e27–9.

5. Simard, J. M., Sheth, K. N., Kimberly, W. T., Stern, B. J., del Zoppo, G. J., Jacobson, S., et al. (2014). Glibenclamide in cerebral ischemia and stroke. Neurocrit. Care 20, 319– 333.

6. Van den Steen, P. E., Dubois, B., Nelissen, I., Rudd, P. M., Dwek, R. A., and Opdenakker, G. (2002). Biochemistry and molecular biology of gelatinase B or matrix metalloproteinase-9 (MMP-9). Crit. Rev. Biochem. Mol. Biol. 37, 375–536. doi: 10.1080/10409230290771546

7. Sternlicht, M. D., and Werb, Z. (2001). How matrix metalloproteinases regulate cell behavior. Annu. Rev. Cell Dev. Biol. 17, 463–516. doi: 10.1146/annurev.cellbio.17.1.463

8. Rosenberg, G. A., and Yang, Y. (2007). Vasogenic edema due to tight junction disruption by matrix metalloproteinases in cerebral ischemia. Neurosurg. Focus 22, E4. doi: 10.3171/foc.2007.22.5.5

9. Fujimura, M., Gasche, Y., Morita-Fujimura, Y., Massengale, J., Kawase, M., and Chan, P. H. (1999). Early appearance of activated matrix metalloproteinase-9 and blood-brain barrier disruption in mice after focal cerebral ischemia and reperfusion. Brain Res. 842, 92–100. doi: 10.1016/S0006-8993(99)01843-0

10. Reynolds, M. A., Kirchick, H. J., Dahlen, J. R., Anderberg, J. M., McPherson, P. H., Nakamura, K. K., et al. (2003). Early biomarkers of stroke. Clin. Chem. 49, 1733–1739. doi: 10.1373/49.10.1733

11. Mun-Bryce, S., and Rosenberg, G. A. (1998). Matrix metalloproteinases in cerebrovascular disease. J. Cereb. Blood Flow Metab. 18, 1163–1172.

12. Li, D. D., Song, J. N., Huang, H., Guo, X. Y., An, J. Y., Zhang, M., et al. (2013). The roles of MMP-9/TIMP-1 in cerebral edema following experimental acute cerebral infarction in rats. Neurosci. Lett. 550, 168–172. doi: 10.1016/j.neulet.2013.06.034

13. Rosell, A., Ortega-Aznar, A., Alvarez-Sabín, J., Fernández-Cadenas, I., Ribó, M., Molina, C. A., et al. (2006). Increased brain expression of matrix metalloproteinase-9 after ischemic and hemorrhagic human stroke. Stroke 37, 1399–1406. doi: 10.1161/01.STR.0000223001.06264.af

14. Inzitari, D., Giusti, B., Nencini, P., Gori, A. M., Nesi, M., Palumbo, V., et al. (2013). MMP9 variation after thrombolysis is associated with hemorrhagic transformation of lesion and death. Stroke 44, 2901–2903.

15. Piccardi, B., Palumbo V., Nesi, M., Nencini, P., Gori, A.M., Giusti, B., et al. (2015). Unbalanced metalloproteinase-9 and tissue inhibitors of metalloproteinases ratios predict hemorrhagic transformation of lesion in ischemic stroke patients treated with thrombolysis: results from the MAGIC study. Front. Neurol. 6:121.

16. Adams HP Jr, Davis PH, Leira EC, Chang KC, Bendixen BH, Clarke WR, et.al. Baseline NIH Stroke Scale score strongly predicts outcome after stroke: A report of the Trial of Org 10172 in Acute Stroke Treatment (TOAST). Neurology. 1999; 13:126–131.

17. Hoda MN, Li W, Ahmad A, Ogbi S, Zemskova MA, Johnson MH, et.al. Sex-independent neuroprotection with minocycline after experimental thromboembolic stroke. Exp Transl Stroke Med 2011; 3:16.

18. Heo JH, Kim SH, Lee KY, Kim EH, Chu CK, Nam JM. Increase in plasma matrix metalloproteinase-9 in acute stroke patients with thrombolysis failure. Stroke 2003; 34:E48–50.

19. Montaner J, Alvarez-Sabín J, Molina CA, Anglés A. Abilleira S, Arenillas J, et.al. Matrix metalloproteinase expression is related to hemorrhagic transformation after cardioembolic stroke. Stroke 2001; 32:2762–2767.

20. Tsuji K, Aoki T, Tejima E, Arai K, Lee SR, Atochin DN, et.al. Tissue plasminogen activator promotes matrix metalloproteinase-9 upregulation after focal cerebral ischemia. Stroke 2005; 36:1954–1959.

21. Kumari R, Willing LB, Patel SD, Baskerville KA, Simpson IA. Increased cerebral matrix metalloprotease-9 activity is associated with compromised recovery in the diabetic db/db mouse following a stroke. J Neurochem 2011; 119:1029–40.

22. Uemura S, Matsushita H, Li W, Glassford AJ, Asagami T, Lee KH, et.al. Diabetes mellitus enhances vascular matrix metalloproteinase activity: role of oxidative stress. Circ Res 2001; 88:1291–1298.

23. Planas AM, Solé S, Justicia C. Expression and activation of matrix metalloproteinase-2 and -9 in rat brain after transient focal cerebral ischemia. Neurobiol Dis 2001; 8:834–846.

24. Montaner J, Alvarez-Sabín J Molina C, Anglés A, Abilleira S, Arenillas J, et.al. Matrix metalloproteinase expression after human cardioembolic stroke: temporal profile and relation to neurological impairment. Stroke 2001; 32:1759–1766.

25. Kaste M, Waltimo O. Prognosis of patients with middle cerebral artery occlusion. Stroke 1976; 7:482–485.

26. Fink JN, Selim MH, Kumar S, Silver B, Linfante I, Caplan LR, et.al. Is the association of National Institutes of Health Stroke Scale scores and acute magnetic resonance imaging stroke volume equal for patients with right-and left-hemisphere ischemic stroke? Stroke 2002; 33:954–958.

27. Bharosay A, Bharosay VV, Varma M, Saxena K, Sodani A, Saxena R. Correlation of Brain Biomarker Neuron Specific Enolase (NSE) with Degree of Disability and Neurological Worsening in Cerebrovascular Stroke. Indian J Clin Biochem 2012; 27:186–190.

28. González-García S, González-Quevedo A, Fernández-Concepción O, Peña-Sánchez M, Menéndez-Saínz C, Hernández-Díaz Z, et.al. Short-term prognostic value of serum neuron specific enolase and S100B in acute stroke patients. Clin Biochem 2012; 45:1302–1307.

29. Kashyap RS, Kabra DP, Nayak AR, Mishra RA, Deshpande SK, Karandikar PN, et.al. Protein electrophoretogram in serum of Acute Ischemic Stroke patients & its correlation with S-100 ßß and Neuron Specific Enolase level: A pilot study. Annals of Neurosciences, 2006, 13; 36–40.

30. Kashyap RS, Nayak AR, Deshpande PS, Kabra DP, Purohit HJ, Taori GM, et.al. Inter-alpha-trypsin inhibitor heavy chain 4 is a novel marker of acute ischemic stroke. Clin Chim Acta 2009; 402:160–163.

